# Metagenomic analysis after selective culture enrichment of wastewater demonstrates increased burden of antibiotic resistant genes in hospitals relative to the community

**DOI:** 10.1101/2023.03.07.23286790

**Authors:** Nicole Acosta, Jangwoo Lee, Maria A. Bautista, Srijak Bhatnagar, Barbara J. Waddell, Emily Au, Puja Pradhan, Rhonda G. Clark, Jon Meddings, Norma Ruecker, Gopal Achari, Johann D. Pitout, John Conly, Kevin Frankowski, Casey R.J. Hubert, Michael D. Parkins

## Abstract

Antimicrobial resistance (AMR) is an ever-increasing threat to global health. Wastewater-based surveillance is an emerging methodology that objectively enables an inclusive and comprehensive assessment of population AMR in an observed sewershed. Here we compared the resistome of two tertiary-care hospitals with two separate neighborhoods, using complimentary targeted qPCR and metagenomics of wastewater before and after selective culture enrichment for clinically important Gram negatives. In total 26 ARG-type (1225 ARG-subtypes) were found across all samples, in which β-lactam ARG was the richest (the number of different ARG-subtypes found) followed by multidrug, fluoroquinolone, macrolide-lincosamide-streptogramin (MLS) and aminoglycoside. The composition of ARGs in wastewater differed between raw wastewater pellets and culture-enriched wastewater samples and the resistomes clustered based on the type of location (Hospitals vs neighborhoods). Hospital wastewater was found to have higher diversity and greater abundance of ARGs compared to neighborhood wastewater when the composition profiles of ARGs in both raw and culture-enriched wastewater pellets. Clinically relevant ARG (i.e., VIM, NDM metallo-ß-lactamases) were detected in culture enrichment samples that were not identified in raw samples, despite a lower targeted sequencing depth. Wastewater-based surveillance is an effective, and potentially extremely important and powerful tool that could be developed to augment hospital-based infection control and antimicrobial stewardship programs, creating a safer space for those receiving care.

## INTRODUCTION

The emergence and spread of antimicrobial-resistant organisms (AROs) is a profound global health threat (Mulvey and Simor 2009; Finaly B et al. 2019). AROs result in disproportionally higher morbidity and mortality, and incur huge healthcare costs (Cosgrove et al. 2003; Tumbarello et al. 2015; Morata et al. 2012; Zimlichman et al. 2013; Valiquette, Chakra, and Laupland 2014; Ghantoji et al. 2010). Globally, ARO-related deaths are expected to rise to 10 million/year by 2050, costing the global economy >$100 trillion in lost productivity (Strathdee, Davies, and Marcelin 2020). In particular, emerging threats such as Enterobacterales and *Pseudomonas aeruginosa*-producing extended-spectrum β-lactamases (ESBLs) (Peirano et al. 2020; Holland et al. 2020; Peirano et al. 2012; Pitout et al. 2007; Pitout et al. 2004), metallo-beta-lactamases (MBLs) (Matsumura et al. 2018; Peirano et al. 2018; Parkins et al. 2007; Pitout, Nordmann, et al. 2005; Pitout, Gregson, et al. 2005; Doyle et al. 2012) and KPC carbapenemases (Doyle et al. 2012; Stoesser et al. 2017; Chan et al. 2013) are associated with increasing incidence, and few potential treatment options and thusly recognized as key organisms of concern by the World Health Organization and US Centers for Disease Control and Prevention.

Recently Mitchell and collaborators (2022) performed an exploratory analysis of AMR and proposed a framework to align AMR research with AMR policy and its connections across the One Health landscape (i.e., humans–animals–environment). Two of the eight key themes that were emphasized was the optimizations of AMR surveillance and the understanding of AMR in the environment (Mitchell, O’Neill, and King 2022). These key aspects require a broad knowledge of resistance burden within the community and understanding of the drivers behind AMR outbreaks/hotspots. In the light of this, wastewater-based surveillance (WBS) has been described as a potential tool that could be adapted for this purpose. Indeed, WBS has proven transformative in understanding the COVID-19 pandemic (Acosta et al. 2021; Acosta et al. 2022; Acosta et al. 2023; Peccia et al. 2020; Randazzo et al. 2020; Miyani et al. 2021; Zhu et al. 2021; Xiao et al. 2021; Zhan et al. 2022; Galani et al. 2022; Hegazy et al. 2022; Hubert et al. 2022). WBS’s major advantage is its ability to objectively monitor entire populations passively and comprehensively, making it accurate, inclusive and cost-effective (Choi 2020; Sims and Kasprzyk-Hordern 2020).

Multiple approaches have been used in initial investigations of AMR in wastewater including both culture-dependant and independent approaches. These works have increasingly identified clinically relevant ARGs in wastewater, including Metallo-β-lactamases (MBLs) (e.g., New Delhi Metallo-beta-lactamases (NDM), VIMs, IMPs) and mobile colistin resistance (i.e., Mcr-1) (Mittal 2019; Lira et al. 2020; Fleres et al. 2019). A limitation of metagenomic assessment of environmental samples relates to sequencing depth, with progressively greater costs incurred with higher sequencing depths when assessing for ultra-rare gene targets (Zhang et al. 2015; Wang et al. 2015). Furthermore, accurate gene identification and classification is dependant on factors including efficient assembly of short-read fragments into large contigs, inhibitory factors in the matrix, and contaminating host DNA. Selective culture enrichment is an elegant technique, uniquely positioned to augment metagenomic assessment of complex samples overcoming some of these limitations (Whelan et al. 2020; Zhang, Zhang, and Ju 2022; Wang et al. 2021). For instance, manipulating growth conditions, specific phenotypes of interest (i.e., those mediate via transmissible beta-lactamases) can be differentially increased potentially improving the detection limits of agnostic approaches for ultra-rare genes. Furthermore, culture enrichment also increases the potential to ascribe individual ARG to those organisms carrying them in the natural environment (Miłobedzka et al. 2022; Marano et al. 2021).

Herein, we assessed the resistome of wastewater from multiple tertiary care hospitals and neighborhoods in Calgary, Canada, by deploying complementary metagenomic analysis of raw pellet and selectively culture-enriched wastewater samples to identify genes mediating antimicrobial resistance of critical pathogens. Hospitals represent a unique and under explored area for wastewater-based surveillance applications. Indeed, infections represent the area where hospitals are most vulnerable, yet least resourced to respond dynamically, and new tools to augment infection control, and antimicrobial stewardship are desperately required. The aims of this work were i). establish if the resistome of hospital wastewater differed from the general population, ii). determine if specific class of ARG existed disproportionally in hospitals and iii). determine if selective culture enrichment would increase the potential of conventional metagenomics approaches in detecting ultra-rare ARG of clinically relevant Gram negatives.

## MATERIAL AND METHODS

### Wastewater collection

Twenty-four-hour composite wastewater was collected from two hospitals (Hospital-1 and Hospital-2) and two neighborhood locations (NE1 and NW1) (Supplementary Figure 1) in the summer of 2021 across Calgary, Alberta Canada. Hospital sampling locations were chosen for their comprehensive and exclusive coverage of each site’s sewershed (Acosta et al. 2021). These two hospitals encompass 47% of regional tertiary-care adult beds in the city of Calgary: Hospital-1 (Northeast Calgary, ∼511 inpatient beds) and Hospital-2 (Southwest Calgary, ∼611 inpatient beds). Neighborhood locations were chosen based on ease of access and similar size of population served (i.e., NE1: 44,839 and NW1: 46,008), where population size was estimated based on the number of service connections (Acosta et al. 2022) and to capture populations of diverse socioeconomic backgrounds. For example, NE1 relative to NW1 had an average household size of 3.78 vs NW1 2.72 individuals; percentage of constituents belonging to a visible minority of 81.1% vs 35.6 and an average household income of $36,809 (Canadian) vs 68,228 as determined using federal census data (ref). Wastewater samples were time-weighted, in which autosamplers were programmed to collect 100 ml of wastewater every 15 mins, for a total of 96 pooled samples over a period of 24 h. Samples were collected and stored on ice by City of Calgary staff using a standardized protocol (Acosta et al. 2021; Acosta et al. 2022). This project was approved by the Conjoint Regional Health Ethics Board (REB20-1252).

### Pellet collection and culture enrichment

Wastewater pelleting was adapted from a published protocol (Che et al. 2019). Briefly, two aliquots of 40 ml were taken from each composite sample after thorough mixing. Samples were centrifuged at 4200 rpm for 20 minutes at 4□. The supernatant from each was carefully removed and discarded, and the pellets combined. The total volume of pellet recovered was recorded and used for further calculations. All processed pellet samples were either stored at −20°C before DNA extraction or were immediately plated for culture-enrichment steps.

In an attempt to increase detection of Gram-negative resistance mechanisms, 50 µl of each fresh pellet was plated and spread onto MacConkey agar plates supplemented with either 8µg/ml meropenem (MEM), 4µg/ml ciprofloxacin (CIP), 8µg/ml ceftriaxone (CRO) or 2µg/ml colistin (COL). Plates were incubated overnight (12 h) at 37 °C. Bacterial growth was collected by adding 1 ml of UltraPure™ DNase/RNase-Free Distilled Water (Invitrogen) to plates, physical scraping to collect growth, and the final volume collected. The total volume of the culture-enriched wastewater pellet recovered was recorded and used for further calculations. All the processed culture-enriched wastewater pellets were stored at −20°C before DNA extraction.

### DNA extraction, library preparation and shotgun metagenomic sequencing

Aliquots of 250 µl of either the raw wastewater pellet or culture-enriched pellet were used for total genomic DNA extraction using the PowerSoil DNA Isolation Kit (Qiagen) according to the manufacturer’s recommendations. An extraction blank control was included for every processed sample batch to ensure no contamination occurred. DNA concentration was measured using the Qubit fluorometer (Thermo Fisher Scientific). All extracted DNA samples were sent out for library preparation and shotgun metagenomic sequencing to Novogene Corporation Inc., California, USA. Paired-end sequencing (2 × 150 bp) sequencing was performed on Illumina Novaseq 6000 PE150 platform with a sequencing output target of ∼10 giga-bases (∼33M paired reads/sample) for the raw pellets and 6 giga-bases (∼20M paired reads/sample) for the culture-enriched samples (Supplementary Table 1).

### Bioinformatics workflow

Illumina raw reads were first filtered by our sequencing provider (Novogene, USA). In particular, i) reads containing adapters were removed, ii) reads containing N>10% were removed, where N represents a base that cannot be determined; and iii) reads containing low quality (Q score ≤ 5) were removed. The base qualities and adapter contamination were double-checked by the authors using FastQC (v.0.11.9)(Andrews 2010), and further trimmed using Trimmomatic (v0.39) (Bolger, Lohse, and Usadel 2014). For annotating ARGs to the trimmed reads, we used DeepARG-SL pipeline v2.0, which aligns reads against DeepARG-DB v2.0 database which includes ARGs found from multiple public databases (i.e., CARD, ARDB, UniProt, and also 16S rRNA gene databases (i.e., SILVA and Greengenes)) (Arango-Argoty et al. 2018; Arango-Argoty et al. 2016). The DeepARG-SL pipeline v2.0 outputs were subject to further analysis, for instance to calculate coverage (or ARG copy) (E1), and normalized ARG abundance (E2) for each subtype homologues (e.g., OXA-1, -2, etc.) according to the customized workflow written in Python (v3.5.3).

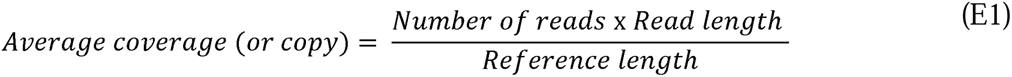

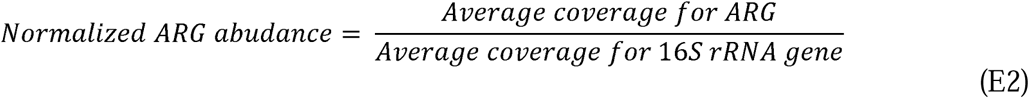

where number of reads is the number of ARG read identified to a specific ARG; read length is the sequence length (bp) of the Illumina reads (i.e., 150 bp); reference length is the sequence length of the corresponding target ARG sequence referring to DeepARG-DB v2.0 database (Arango-Argoty et al. 2018); Number of 16S sRNA copies is the number of the 16S rRNA gene sequence identified in each sample; and the 16S rRNA gene length is the sequence length of 16S rRNA gene (i.e., 1432 bp) (Yang et al. 2016). All downstream analyses was performed using 16S rRNAs-normalized ARG abundance hereafter. Identified ARG subtypes were categorized and aggregated by ARG-types which refers to the antimicrobial agents to which each gene confers resistance (Figure 1A). The workflow using an example dataset will be available in the second author’s GitHub page (https://github.com/myjackson) upon acceptance of this manuscript to a journal or request from reviewer. The raw reads will be available in the National Center for Biotechnology Information (NCBI) Sequence Read Archive (SRA) repository, under the BioProject ID XXXX (http://www.ncbi.nlm.nih.gov/bioproject/XXXX). The data will be released to the public when the manuscript is formally accepted for publication.

**Figure 1.**
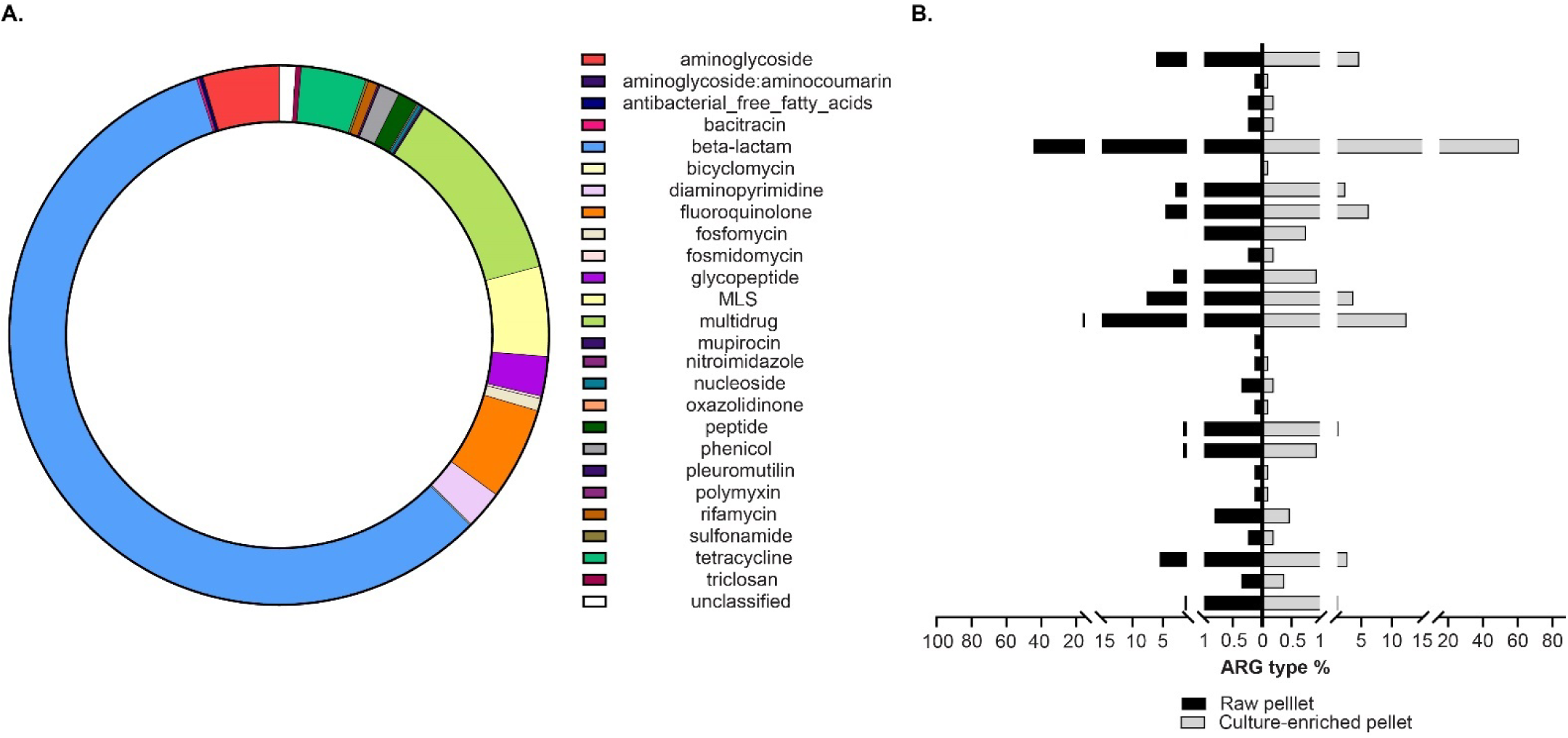
Composition profile of ARG-types in wastewater. **A.** Percentage of the resistome for each ARG-types (mechanisms for antibiotic resistance) category found in samples before and after culture enrichment from hospital and neighborhoods locations. **B.** ARG-type percentage classified by the type of sample (i.e., Raw wastewater pellet and culture-enriched wastewater pellet).

### ARG qPCR assays

qPCR was performed to objectively quantify the absolute abundance of different ARGs of interest (i.e., *bla*_CTX-M-1_, *bla*_KPC-3_, *bla*_NDM-1_, *vanA*) in raw and enriched wastewater pellets. For each ARG detection, 10 µl reaction contained 5 µl of TaqMan® Fast Advanced Master Mix (Applied Biosystems), 0.5 µl of TaqMan Gene Expression assay (Thermo Fisher Scientific) (Supplementary Table 2), and 4 µl of the template DNA. For all ARG qPCR assays, the qPCR program reaction consisted of an initial step at 50°C for 2 min, then a step at 95°C for 2 min followed by 40 cycles of 95°C for 1 s, 60°C for 20 s. A 9-fold dilution series of double-stranded DNA fragments (Integrated DNA Technologies (IDT), USA) were synthesized for each ARO qPCR assay (Supplementary Table 3) and were run in triplicate on every 96-well PCR plate to produce standard curves and used to estimate the absolute quantity (copies/µl).

For total burden of specific antimicrobial resistant organisms (AROs) and associated determinants (i.e., *Clostridioides difficile* (CD) and *tcdA*), each 10 µl reaction contained 5 µl of TaqMan® Fast Advanced Master Mix (Applied Biosystems), 0.5 µl of a mix of forward and reverse primers, 0.5 µl of probe (Supplementary Table 2); and 4 µl of the DNA template. For both CD 16S rRNA and *tcdA* qPCR assays, the qPCR program reaction consisted of an initial step at 50°C for 2 min, then a step at 95°C for 2 min followed by 50 cycles of 95°C for 1 s, 56°C for 20 s. A 7-fold dilution series of double-stranded DNA fragments (Integrated DNA Technologies (IDT), USA) was synthesized and used (Supplementary Table 3) and were run in triplicate on every 96-well PCR plate to produce standard curves and used to estimate the absolute quantity (copies/µl).

For total bacterial load determination (16S rRNA gene copies/ml of wastewater), each 10 µl reaction contains 5 µl of TaqMan® Fast Advanced Master Mix (Applied Biosystems), 0.5 µl of a mix of forward and reverse primers, 0.5 µl of probe (Supplementary Table2); and 2.5 µl of the DNA template. The qPCR program reaction consisted of an initial step at 50°C for 2 min, then a step at 95°C for 2 min followed by 40 cycles of 95°C for 1 s, 62 °C for 20 s. A 9-fold dilution series of *P. aeruginosa* genomic DNA PA01 strain (62.5 to 1.6 × 10^-4^ ng/µl) was used as standard curve. Data were converted from DNA concentration (ng/µl) to 16S rRNA copy number as previously described by Zhao et al (Zhao et al. 2012). The number of 16S copies in a given reaction was multiplied by the total number of nanograms in the entire sample DNA prep and total eluted volume after DNA extraction and then divided by the volume of wastewater used for DNA extraction to give an absolute abundance of 16S copies/ml of wastewater. All samples were assayed in duplicate, and all runs contained non-template control. All qPCR reactions were carried out in the QuantStudio 5 Real-time PCR systems (Applied Biosystems).

### Statistical analysis

Alpha-diversity was calculated to estimate the ARG diversity from the metagenomic sequencing data using Shannon Diversity index (SDI). SDI was compared between hospitals (n = 8) and neighborhoods (n=8) locations from the culture-enriched wastewater pellet samples using Mann-Whitney test (α = 0.05). Kruskal-Wallis Test was performed to test for differences in SDI across samples after culture-enrichment based on the antibiotics used. Beta-diversity clustering plots (i.e., non-metric multidimensional scaling (NMDS)) were generated using the Bray-Curtis dissimilarities metrics of the ARG-subtypes to visualise potential clustering patterns among samples. Analysis of similarity (ANOSIM) (Somerfield, Clarke, and Gorley 2021) was conducted to determine the significance of dissimilarities between/among groups. Permutational multivariate analysis of variance (PERMANOVA) tests were performed to test differences in the Bray-Curtis distance matrices. Both ANOSIM and PERMANOVA were performed with permutations=999 using the vegan package in R. Two-proportions z-test was used to evaluate the richness distribution of ARG-types across all samples. When appropriate, P-values were adjusted for multiple comparisons by using Benjamini–Hochberg P-value correction for multiple testing. All data analyses and visualizations were performed in GraphPad Prism-9 software (La Jolla, CA) and in R (V4.0.4).

## RESULTS

### General composition profiles of ARGs in wastewater samples

Metagenomic DNA isolated from wastewater samples was used to determine ARGs profiles. Shotgun metagenomic sequencing yielded 1.24 billion raw reads; 31.8% and 68.2% of those sequences were obtained from raw wastewater pellets (n= 4) and culture-enrichment samples (n=16), respectively. Sequencing produced (mean ± SD) ∼98.6 ± 22.1 million raw reads per raw wastewater pellet, and 49.7 ± 13.7 million reads per culture-enriched pellet sample (Supplementary Table 1). Out of the total sequenced reads, 5.3 million raw reads were successfully aligned to the DeepARG-database and used for the normalization analysis. To compare levels of ARGs between groups we normalized the abundance of each ARG by dividing raw reads by the total abundance of the 16S rRNA copies. ARGs in the all samples comprised 26 main ARG-types (Figure 1A) and 1225 ARG-subtypes (Supplementary Table 4), of which the most rich (the number of different ARG-subtypes found) ARG-type that was identified was β-lactam resistance (57.6%), followed by multidrug (11.8%), fluoroquinolone (5.6%), macrolide-lincosamide-streptogramin (MLS) (5.4%), aminoglycoside (4.6%), tetracycline (3.9%) and glycopeptide (2.4%) resistance genes (Figure 1A). There were differences in the distribution of the proportion of some ARG-type identified between pellet and culture-enrichment samples (Figure 1B). The proportion of β-lactam ARGs-type identified in samples selectively enriched by culture was greater than in raw pellets (adjusted P-value <0.05). In contrast, multidrug, MLS, tetracycline and glycopeptide ARGs-types identified in raw wastewater pellets was greater than in culture enriched samples (adjusted P-value <0.05).

### Composition profiles of ARGs in wastewater differs between raw wastewater pellet and culture-enriched wastewater pellet

To determine whether culture enrichment was a suitable approach for identifying ARG in wastewater samples, we calculated a normalized abundance ratio of a given ARG-type after culture enrichment relative to raw pellets (Enrichment factor (EF)). Out of the 26 ARG-types found in this study (Figure 1), 19 (73%) were found to have increased abundance compared to raw wastewater pellets regardless of the antibiotic that was used for selection (Table 1). The ARG-types categories which had the greatest increase after culture enrichment was polymyxin, (median EF of 23.9, IQR 12.1-37.9) followed by aminoglycoside:aminocoumarin, nitroimidazole, and triclosan. In contrast tetracycline, bacitracin and rifamycin had the smallest EF across all categories. A few ARG-types were found to have EF decrease after culture enrichment regardless of conditions tested (i.e., glycopeptide) or in neighborhood samples regardless of the antibiotic that was used for selection (i.e., antibacterial free fatty acids, and pleuromutilin) (Table 1). Bicyclomycin was the only ARG-type that was detected only in culture-enriched wastewater pellets but not in wastewater pellet samples.

**Table 1.**
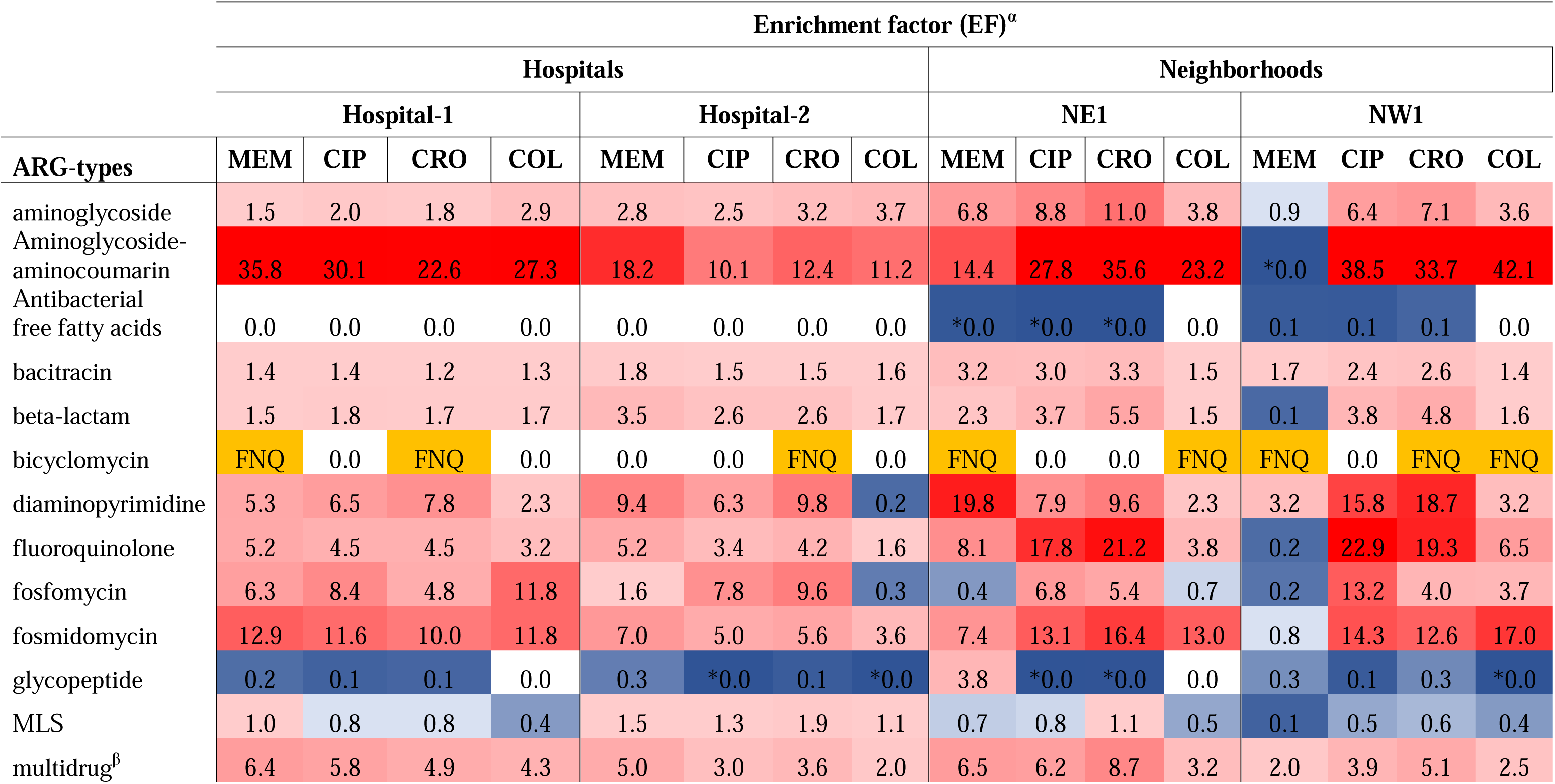
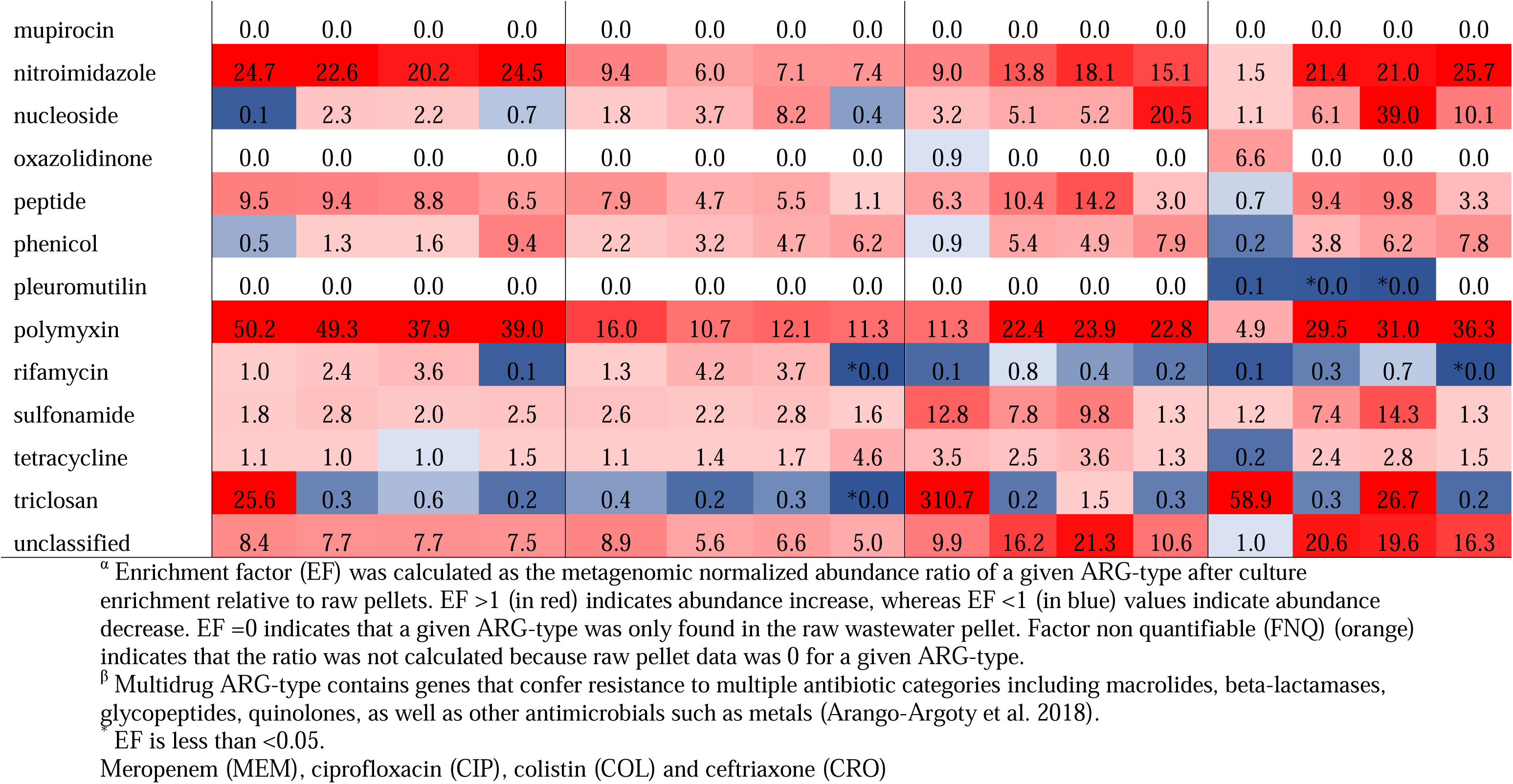
Changes in ARG-types after culture-enrichment of wastewater on MacConkey agar plates supplemented with selective antibiotics relative to pelleted wastewater.

To better understand the differences in resistomes across locations and conditions, we calculated the Bray-Curtis dissimilarities and found a clustering of samples based on treatment (i.e., before and after culture enrichment) and sample location (i.e., hospital and neighborhood) (Figure 2). We performed ANOSIM test and found that overall beta diversity values by treatment (ANOSIM statistic R=0.799, P=0.0012) and sample location (ANOSIM statistic R=0.112, P=0.0463) were significantly different. To further test the effect of treatment and sample location we performed PERMANOVA test to determine the effect of those variable on the resistome. ARG-subtypes profiles were found to be distinguishable by treatment (R^2^=41.7, P=0.001) but not by collection location (R^2^=9.6, P=0.09).

**Figure 2.**
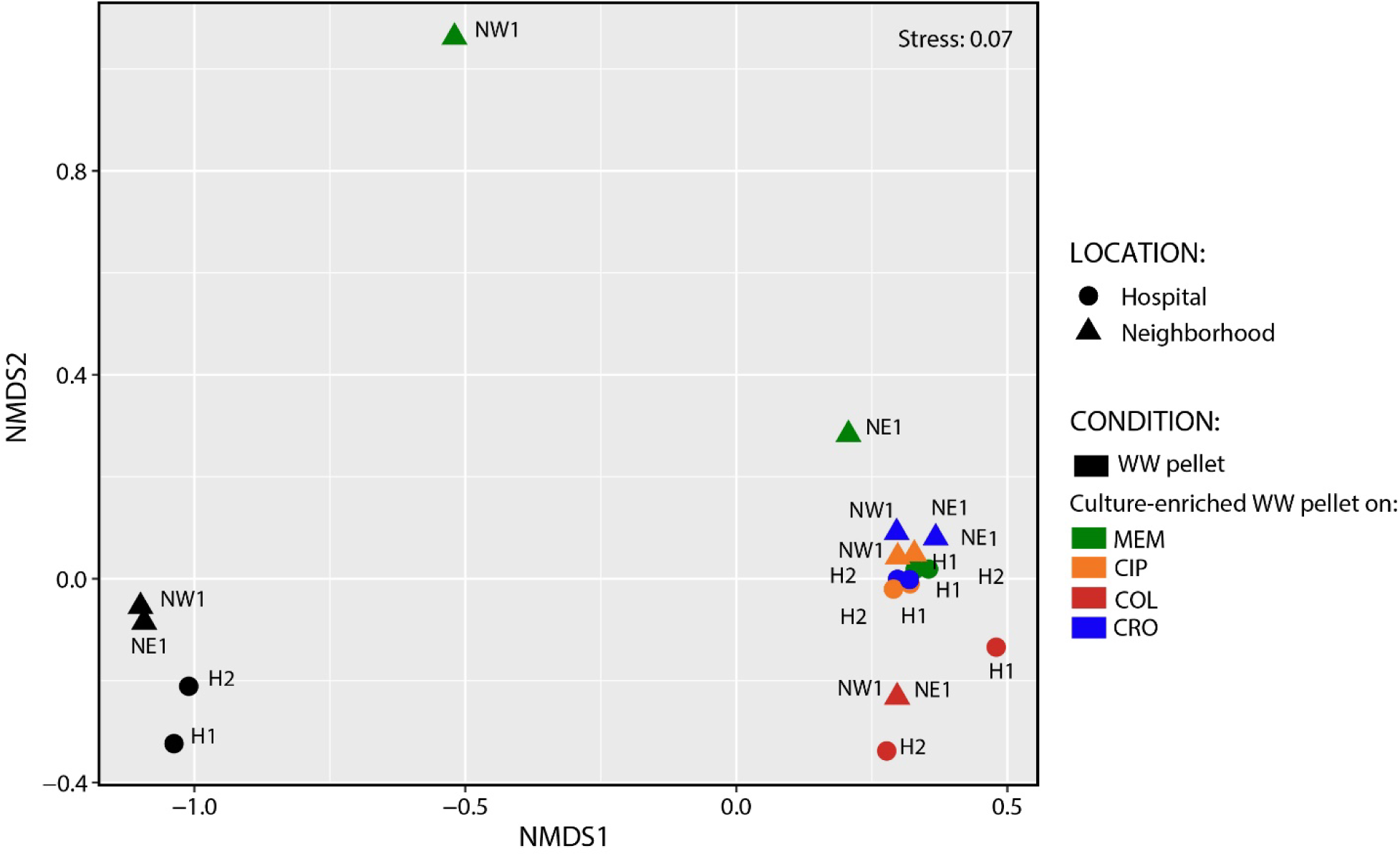
Non-metric multidimensional scaling (NMDS) plot for the distribution of wastewater ARG-subtypes before and after culture enrichment. NMDS were done on the Bray-Curtis distances of ARG profiles. Samples were shape-coded based on the location where wastewater was collected (i.e., Hospital or neighborhood); and color-coded based on condition analysed (i.e., raw pellet versus culture-enriched pellet). H1: Hospital-1, H2: Hospital-2, MEM: meropenem, CIP: ciprofloxacin, COL: colistin and CRO: ceftriaxone.

Because β-lactam resistance was identified as the most diverse ARG-type and its clinical importance, we further analysed the normalized abundance ratio of specific β-lactamases before and after culture enrichment. β-lactamases were grouped by the Ambler classification and the EF was calculated. For most of the β-lactamases analysed there was an increase in their abundance after enrichment (Figure 3). CTX-M and AmpC had the highest average EF value regardless of the antibiotic used (101 and 90 EF, respectively). The overall average EF values were the smallest for KPC (6.7) and NDM (3.2) across all conditions (i.e., antibiotic selection) (Figure 3). However, KPC and NDM were the least abundant of these gene types, and their EF ratios for many neighborhood samples could not calculated (indicated as FNQ) as they were only identified after enrichment (i.e., not in raw pellet). A few β-lactamases (i.e., OXA and GES) were found to have decreased in abundance after culture enrichment. VIM was only detected at Hospital-1 location, where the EF value was 36.3 with MEM.

**Figure 3.**
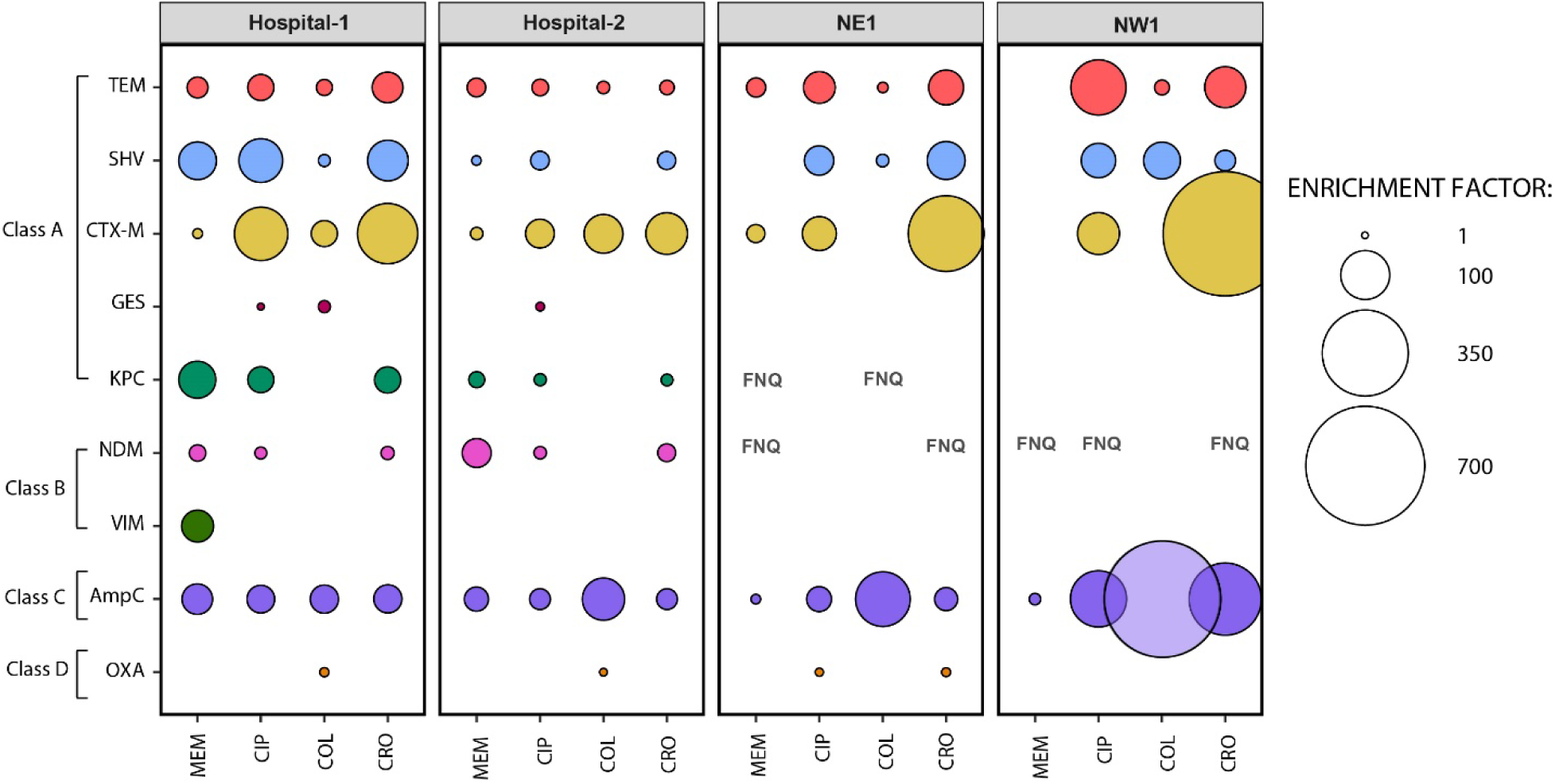
Relative enrichment of β-lactamases found in the wastewater resistome after selective culture-enrichment. Enrichment factor (EF) (shown as bubbles) was calculated as the metagenomic abundance ratio of a given β-lactamase:16S rRNA following growth on MacConkey agar plates impregnated with antibiotics relative to raw wastewater pellets. Circles are shown only when EF > 1. Factor non quantifiable (FNQ) indicate that the ratio was not calculable because raw wastewater pellet contained 0 reads for a given β-lactamase. MEM: meropenem, CIP: ciprofloxacin, COL: colistin and CRO: ceftriaxone.

To characterize in greater detail several of the β-lactamases that we observed EF values increase (i.e., CTX-M and NDM) or decrease (i.e., OXA) after culture enrichment, the normalized ARG abundance was analyzed by ARG-subtypes. We found 61 different CTX-M enzyme types, where CTX-M-90, 29 and 84 were the most abundant, found across all sample locations and antibiotic tested (Figure 4A). CTX-M 14 and CTX-M-15 variants were found in many conditions (i.e., before and after culture enrichment) across all sample locations (Supplementary Table 4). We found 10 NDM variants across all resistomes analyzed. NDM-3, 17 and 2 were the most abundant variants found across all samples (Figure 4B). Finally, we found 152 OXA enzyme variants, where OXA-1, -3 and -9 were the most abundant across all samples (Figure 4C).

**Figure 4.**
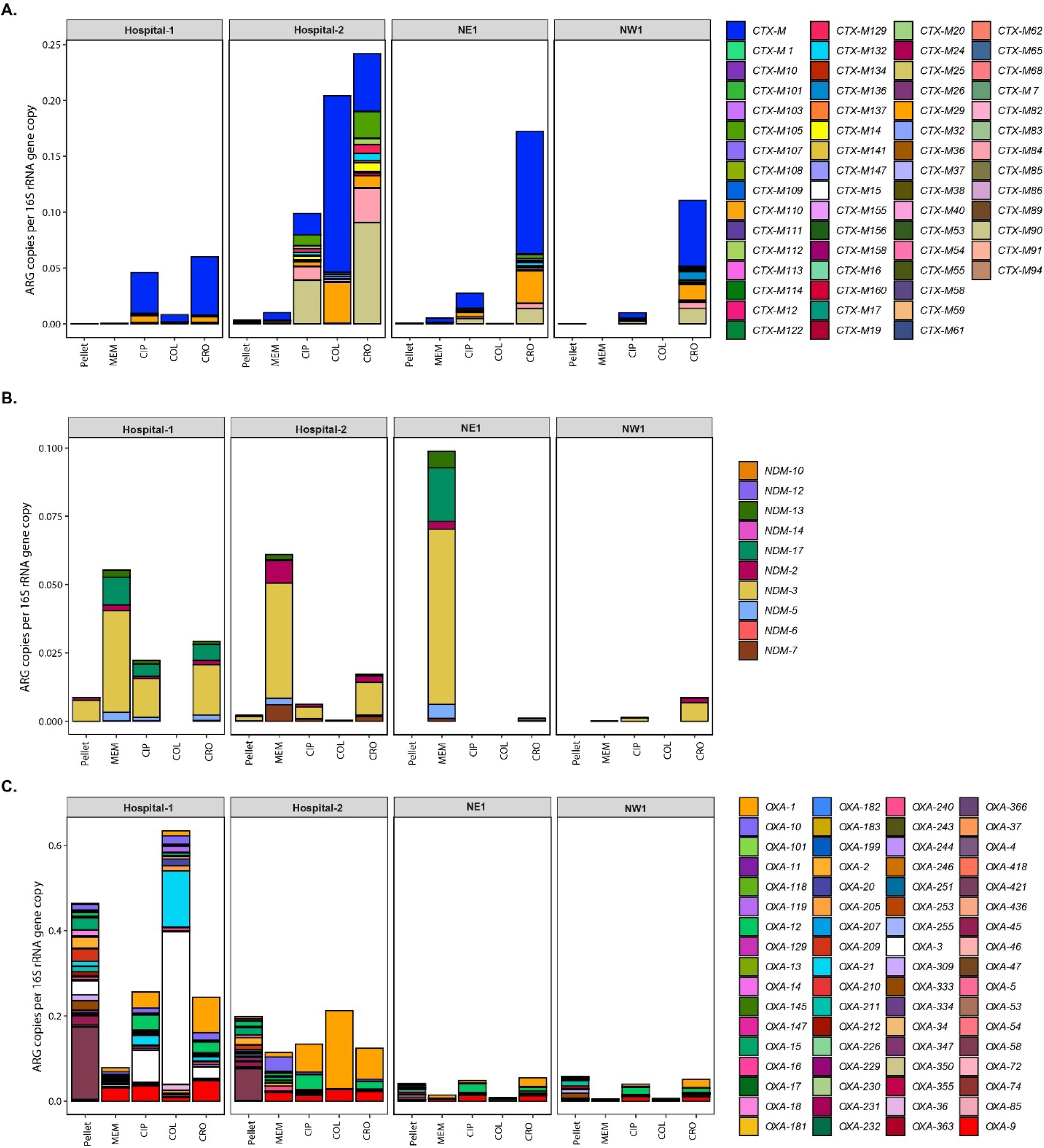
Normalized abundance of clinically importantβ-lactamases found in the resistome of wastewater before and after culture enrichment. Normalized abundance is expressed as ARG copies per 16S rRNA gene copy numbers of CTX-M (**A**), NDM (**B**) and OXA (**C**) from raw wastewater pellet and culture-enriched pellet collected from hospitals (Hospital-1 and Hospital-2) and neighborhoods (NE1 and NW1). ARGs-subtype allelic variants are color-coded. In panel A, CTX-M allelic variants that were not classified as a specific variant during the read based annotation step were cluster and color-coded in the same group. In panel C, for ease of illustration only the 68 out of 152 variants that were found with a normalized abundance >0.0005 within the OXA group were plotted. MEM: meropenem, CIP: ciprofloxacin, COL: colistin and CRO: ceftriaxone.

### Composition profiles of ARGs in culture-enriched wastewater pellet samples

ARG diversity analysis of hospital and neighbourhood wastewater from culture-enriched pellet samples was performed by measuring the Shannon diversity index (SDI). The median SDI for hospitals: 4.7 (IQR: 4.4 - 4.81) and neighborhoods: 4.43 (IQR: 3.91 - 4.54)) was significantly different between locations regardless of enrichment condition (Figure 5A, P< 0.05, Mann-Whitney test). We also analyzed the SDI to understand overall alpha-diversity across all culture-enriched conditions (Figure 5B). Median SDI was highest for wastewater enriched on MacConkey agar plates supplemented with ceftriaxone (SDI median 4.7, IQR: 4.4-4.7), followed by ciprofloxacin (SDI median 4.6, IQR: 4.4-4.7), meropenem (SDI median 4.5, IQR: 3.9-4.6) and colistin (SDI median 3.9, IQR: 3.9-4.2) (P<0.05, Kruskal-Wallis test).

**Figure 5.**
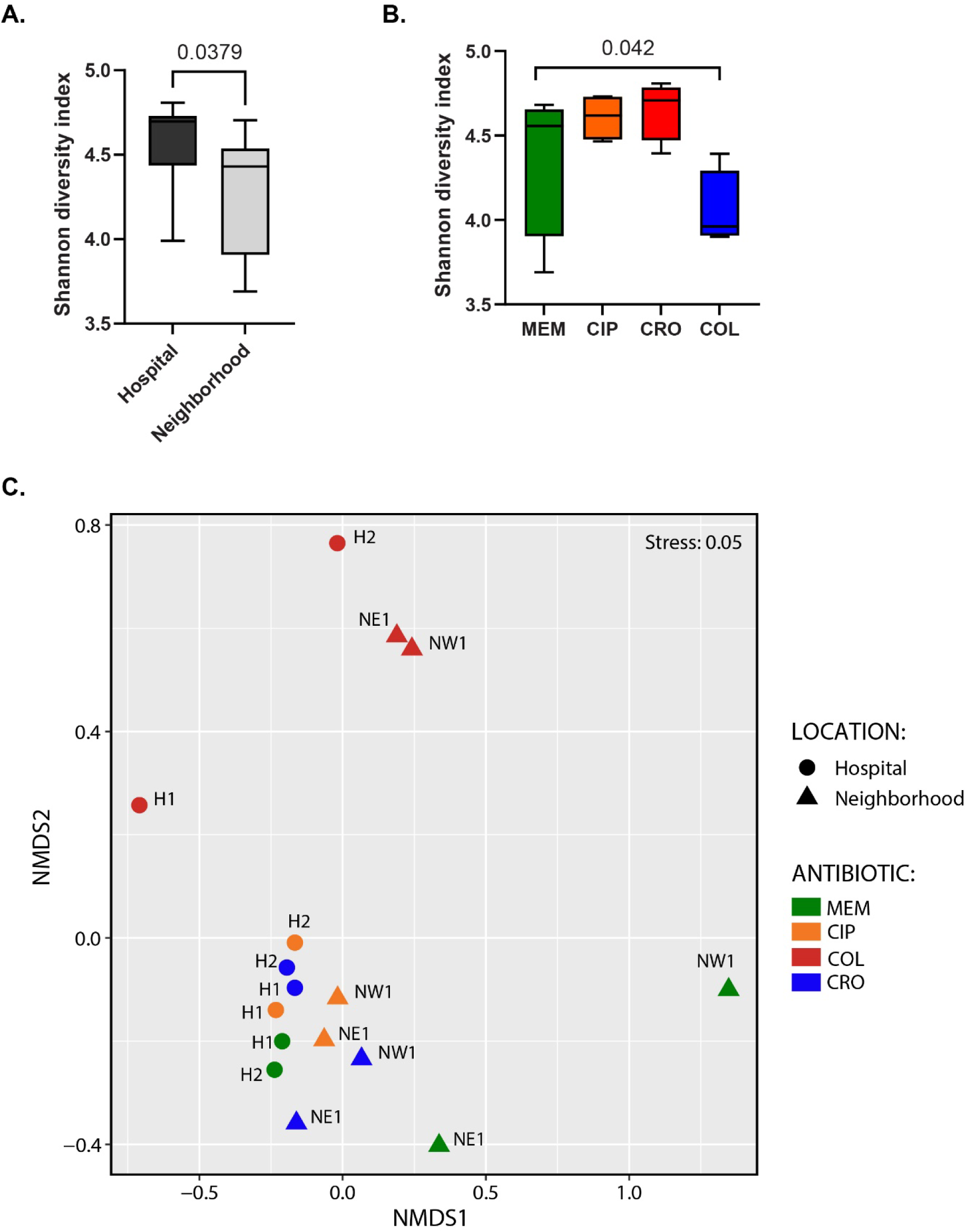
ARGs-subtype alpha and beta diversity of wastewater resistome after culture enrichment. Comparison of the ARG alpha diversity measured using Shannon Diversity index of the culture-enriched wastewater pellet samples based on; (**A**) sample location and (**B**). antibiotic used for selective culture enrichment (i.e., MEM: meropenem, CIP: ciprofloxacin, COL: colistin and CRO: ceftriaxone) as determined by Mann-Whitney test and Kruskal-Wallis Test, respectively. **C**. NMDS plot was done on the Bray-Curtis distances of ARG-subtype profiles of the culture-enriched wastewater pellet samples. Samples were shape-coded based on the type of location where the sample was collected (i.e., Hospital or neighborhood); and color-coded based on the antibiotic used for selection.

Clustering of wastewater resistomes in NDMS ordination plot was shown to be influenced by location sample location as demonstrated by the Ordination plot at the ARG-subtype level (Figure 5C) with similar results at ARG-type (data not shown). Similarly, NMDS ordination plot showed a separation of resistomes based on the antibiotic used for selection. The separation between hospital and neighborhoods and between antibiotic used after culture enrichment was further tested with ANOSIM (ANOSIM statistic R=0.187, P=0.011 and ANOSIM statistic R=0.252, P=0.004 for sample location and antibiotic analyses, respectively) and PERMANOVA tests (R^2^=16, P=0.015 and R^2^=40.03, P=0.004 for sample location and antibiotic analyses, respectively).

Few ARG-types were identified exclusively in the resistome of raw wastewater pellets. Of those, mupirocin was the only ARG-type whose presence was only observed for all conditions and locations tested (Table 1). However, its normalized abundance was amongst the lowest of ARG-types in raw pellets (7.1 x 10^-5^ vs 1.4 x 10^-1^). Similarly, oxazolidinone was only found in wastewater pellet samples with the exception that it was also found in meropenem culture-enriched wastewater pellet samples in both neighborhood locations (i.e., NE1 and NW1) (Table 1). Pleuromutilin and antibacterial free fatty acids ARG-types were also only detected in pelleted samples in only hospital locations (i.e., Hopsital-1 and 2) (Table 1).

### Quantitative confirmation of differential detection of target ARG

We used qPCR to objectively and independently document the abundance of ARG in wastewater pellets. As β-lactam resistance was the most abundant ARG-type that was identified (57.6%), we quantified the absolute abundance of target β-lactamases in raw wastewater pellet samples relative to the total bacterial load (i.e., quantified by the 16S rRNA gene copies). Here we confirmed metagenomic observations (Supplementary Table 4) and established that *bla*_CTX-M-1_, *bla*_KPC-3_ and *bla*_NDM-1_ were 9.6-fold, 4066-fold and 5924-fold higher in raw hospital wastewater pellets compared to neighbourhood samples (P<0.05) (Supplementary Figure 2A). We likewise quantified the presence of *vanA* as a surrogate for Vancomycin-Resistant Enterococci (VRE), total-*C. difficile*, and one of its enterotoxins encoded by the *tcdA* gene in pelleted raw wastewater. We found that *vanA*, total-*C. difficile* and *tcdA* relative abundance were 14.5, 11.8, 19.7 fold-higher in hospital than in neighborhood sites (Supplementary Figure 2B) (P<0.05). Finally, since the culture enrichment of wastewater pellet samples was performed on media (i.e., MacConkey agar plates) that selects against Gram positive bacteria, we decided to confirm by qPCR that *vanA* in culture-enriched wastewater pellet samples was negatively selected. We found a negative selection against the presence of *vanA* gene in MacConkey enriched wastewater relative to raw pellets (Supplementary Figure 2C).

## DISCUSSION

Interest in wastewater-based surveillance (WBS) of ARO has been progressively growing. WBS provides an integrated view of ARG burden across a population that is objective, comprehensive and inclusive. Furthermore, it can be performed serially, and at a tiny fraction of the cost of serial active surveillance to monitor for changes in the prevalence of specific ARG, as might occur in the context of an outbreak (Harrington et al. 2022; Li et al. 2022). Despite the great potential of this technology, its application to hospitals has been limited to date. Hospitals are where societies sickest and most vulnerable are cohorted together to receive care. As a consequence, patients in hospital are simultaneously the most susceptible to acquiring infection and the most likely to experience adverse outcomes as a result. As 37-44% of admitted patients receive antibiotics during their hospital stay, the selection pressure for ARO colonization and subsequent infection is particularly intense and dynamic surveillance mechanisms to monitor for changing prevalence of organisms urgently required to augment antimicrobial stewardship and infection control programs (Taylor et al. 2015; Tan et al. 2017; Rudnick et al. 2020; Hocquet, Muller, and Bertrand 2016). Importantly, this work established that the resistome of hospitals differed significantly from community derived samples, and ARG of notable nosocomial pathogens were markedly increased.

Several early approaches have been used for the analysis of AMR in hospital wastewater. For example, Buelow and collaborators (2018) studied the resistome hospital wastewater compared to receiving urban wastewater but identified only a few ARGs using a high-throughput qPCR (Buelow et al. 2018). Other studies have only been focused on the characterization of the diversity of specific ARGs (e.g., ESBLs and KPCs) in hospital wastewater using culture dependent methodologies (Adekanmbi et al. 2020; Chagas et al. 2011). Culture based studies have identified carbapenemase producing organisms both exclusively in WWTP receiving wastewater from hospitals (Ludden et al. 2017) or across all monitored sites irrespective of contributing hospitals (Blaak et al. 2021). Early metagenomic approaches have been described using hospital wastewater (Rodriguez-Mozaz et al. 2015; Petrovich et al. 2020; Duong et al. 2008; Fuentefria, Ferreira, and Corcao 2011; Szekeres et al. 2017; Hutinel, Larsson, and Flach 2022; Hocquet, Muller, and Bertrand 2016) however, no study has utilized culture-enrichment to study in more greater detail the resistome of complex samples such as hospital wastewater samples. Furthermore, by comparing resistomes from multiple hospitals and neighborhoods, we have increased the robustness of these observations.

Using complementary metagenomic and culture enrichment we identified the presence of ARG encoding resistance to β-lactam, multidrug, fluoroquinolone, MLS, aminoglycoside, tetracycline, glycopeptide, diaminopyrimidine, phenicol, fosfomycin, and others. The most diverse ARGs-types detected conferred resistance to β-lactam in both hospital and neighborhood wastewater. We found that the alpha diversity of ARGs found after cultured enrichment were higher in hospital samples compared to neighborhoods. Similar to our results, a previous study showed that hospital wastewater was found to be richer in ARGs than the other type of samples (e.g., urban wastewater), particularly when comparing ARGs-types related with β-lactam and vancomycin resistance genes (∼15-fold and ∼175 fold higher in hospital wastewater, respectively) (Buelow et al. 2018). These results are also supported by previous studies that have demonstrated that hospital wastewater contains richer and high abundances of ARGs than municipal which has been extrapolated to relate to higher burden of ARG colonization amongst hospitalized individuals (Petrovich et al. 2020; Paulus et al. 2019). Even in the “environmental” One Health compartment, the high concentrations of antibiotics in hospital wastewater relative to municipal wastewater create selective pressure on the microbiota driving resistance (Szekeres et al. 2017). Hospital wastewater therefore serve as a particular hotpot for ARGs and AROs dissemination across the watershed, into the environment and ultimately into humans and animals (Kaur, Yadav, and Tyagi 2020; Rodriguez-Mozaz et al. 2015; Rizzo et al. 2013; Vlahović-Palčevski et al. 2007; Robert et al. 2012). Cai et al. (2021), performed weekly WBS of a single ∼250 bed hospital in Shantou, China for ARG using metagenomics over a period of several months. They compared their wastewater derived data with pathogens from 104 individuals (16% of submitted clinical specimens) and used network analysis to demonstrate strong correlations with many ARG-types and recovered clinical pathogens (Cai et al. 2021).

Although metagenomic sequencing is a robust approach for ARGs identification and characterization relative to traditional targeted studies, next-generation sequencing (NGS) requires tremendous sequencing depth in order to effectively identify AMR due to its low sensitivity and lower limit of detection (Lanza et al. 2018; Guitor et al. 2019). Coupling limited selective culture-enrichment of wastewater with metagenomic sequencing allowed us to detect a range of ARG sub-types (including clinically important KPCs, NDMs and VIM metallo-beta-lactamases) that would otherwise not have been detected, despite a sequencing depth less than half of the raw pellets. Such an approach can manifest in significant cost savings – particularly relevant for high volume longitudinal monitoring programs. Interestingly, increased detection of these targets occurred using ciprofloxacin in addition to both the beta-lactams; meropenem and ceftriaxone, likely relating to the co-location of ARG on transmissible mobile elements in resistant pathogens (Parkins et al. 2007). The exceptional enrichment agent was colistin, which negatively selected against carbapenemases, and only modestly enriched for *mcr*-1 which was exceedingly infrequent in the samples collected (NE-1 and NW-1). We identified bicyclomycin ARG, uniquely in the culture-enriched wastewater pellet samples. Antibiotic resistant to bicyclomycin is mediated by a transmembrane protein that excludes it from penetrating the cell (Malik et al. 2014) and it has been found in *P. aeruginosa* strains responsible for outbreaks (Fonseca et al. 2015).

A common critique of metagenomic studies is the inability to distinguish between live and dead organisms, particularly relevant if infection control measures are to be implemented based on wastewater data. Culture enrichment negates this. Only a few studies have used a similar approach of culture enrichment, but they have only focused on AMR in influent and effluent municipal wastewater rather than hospitals. Zhang and collaborators (2022) assessed the resistome through culture enriched phenotypic metagenomics in municipal wastewater treatment plant and its receiving river samples. This study detected a greater diversity and number of ARGs when coupling culture enrichment and metagenomic analysis than direct metagenomic sequencing, with a particular focus on carbapenemase and ESBLs (Zhang, Zhang, and Ju 2022). Marano and collaborators (2021) used culture-enrichment under copiotrophic conditions to enrich and determine the microbiome and resistomes of AROs and ARGs in soil that was irrigated with treated-wastewater. They observed that cephalosporin and carbapenem resistant *Enterobacterales* only after culture enrichment and not in raw soil samples (Marano et al. 2021). Other alternatives to culture enrichment prior to metagenomic analysis consist of using commercially available AMR target enrichment NGS panels (Harrington et al. 2022).

Targeted surveillance of specific clinically relevant ARG serves to complement metagenomic assessments. Indeed, others have suggested that this approach has increased sensitivity relative to shot gun metagenomics (Miłobedzka et al. 2022). Using this approach, we demonstrated marked increase in clinically relevant ARG in hospital wastewater pellets relative to community samples including β-lactamases from nosocomial Gram negatives (i.e., CTX-M ESBLs, KPCs and NDM) and Gram-positive pathogens (i.e., total *C. difficile* and *tcdA*, and *vanA* from VRE).

This study has several notable limitations. We specifically used a lower sequencing target depth in culture enrichment samples in order to assess if this process could manifest in cost savings for identification of ultra-rare ARG. Owing to this difference, we were unable able to perform direct comparisons in terms of SDI between these data sets. The use of selective culture enrichment, designed to increase the identification of any particular target, will conversely mask alternates. As our goal was to assess for ultra-rare carpabenemases presumed to colonize a tiny fraction of the population where it is a minor constituent within resident microbiota, we enriched with MacConkey agar and appropriate antibiotics to select for these mechanisms. In doing so, we selected against other clinically relevant nosocomial pathogens (i.e., VRE and other Gram positives) which were however readily identified owing to their increased prevalence and abundance in colonized/infected individuals. This study did not assess the relationship between antibiotics released into hospital wastewater and abundance of ARGs found in those samples. Only a few studies have looked at that matter and it has been found a correlation between antibiotic concentrations and resistance in wastewater (Rodriguez-Mozaz et al. 2015; Gao, Munir, and Xagoraraki 2012). Finally, we did not identify the taxonomic identity of those organisms carrying ARG, nor did we assess the role of mobile genetic elements in distribution of antimicrobial resistance in hospital and community samples. However, culture-based enrichment offers the potential to retrospectively go back and attempt to recover individual pathogens carrying an ARG-type of interest for greater characterization through whole genome sequencing analysis.

## Conclusion

Here we were able to demonstrate using multiple independent sampling locations and complementary modalities (i.e., metagenomics and qPCR) that the resistome of hospitals, differs from communities, and is particularly enriched for beta-lactam resistance and nosocomial pathogens such as VRE and *C. difficile*. By adopting a simple, semi-selective culture enrichment step we were able to identify ultra-rare, and clinically relevant ARG such as NDM and KPC while simultaneously reducing the costs associated with sequencing. Wastewater-based surveillance is a potentially extremely important and powerful tool that warrants exploration as a tool to augment hospital-based infection control and antimicrobial stewardship programs.

## Data Availability

All data produced in the present study will be made available upon reasonable request to the authors after it is published

## ACKNOWLEDGMENTS

The investigators are grateful to the staff of the City of Calgary Water Services for their support in providing wastewater samples for assessment.

## SUPPLEMENTARY TABLES

**Table 1S.** Overview of the paired-end sequencing (2 × 150 bp) sequencing results obtained by Illumina Novaseq 6000 PE150 platform.

| Sample ID | Output (raw data/sample) | Raw reads | Location | Type |
| --- | --- | --- | --- | --- |
| NA_1 | 10G | 116338888 | Hospital-1 | Pellet |
| NA_2 | 10G | 118220046 | Hospital-2 | Pellet |
| NA_3 | 6G | 57521818 | Hospital-1 | culture-enriched |
| NA_4 | 6G | 49324432 | Hospital-1 | culture-enriched |
| NA_5 | 6G | 57428492 | Hospital-1 | culture-enriched |
| NA_6 | 6G | 49060778 | Hospital-1 | culture-enriched |
| NA_7 | 6G | 49983644 | Hospital-2 | culture-enriched |
| NA_8 | 6G | 53320788 | Hospital-2 | culture-enriched |
| NA_9 | 6G | 59970976 | Hospital-2 | culture-enriched |
| NA_10 | 6G | 40860274 | Hospital-2 | culture-enriched |
| NA_11 | 10G | 74092986 | NE1 | Pellet |
| NA_12 | 10G | 85855692 | NW1 | Pellet |
| NA_13 | 6G | 57157698 | NE1 | culture-enriched |
| NA_14 | 6G | 51630126 | NE1 | culture-enriched |
| NA_15 | 6G | 55454866 | NE1 | culture-enriched |
| NA_16 | 6G | 39933826 | NE1 | culture-enriched |
| NA_17 | 6G | 77036096 | NW1 | culture-enriched |
| NA_18 | 6G | 45107562 | NW1 | culture-enriched |
| NA_19 | 6G | 53285664 | NW1 | culture-enriched |
| NA_20 | 6G | 47359372 | NW1 | culture-enriched |

**Table 2S.**
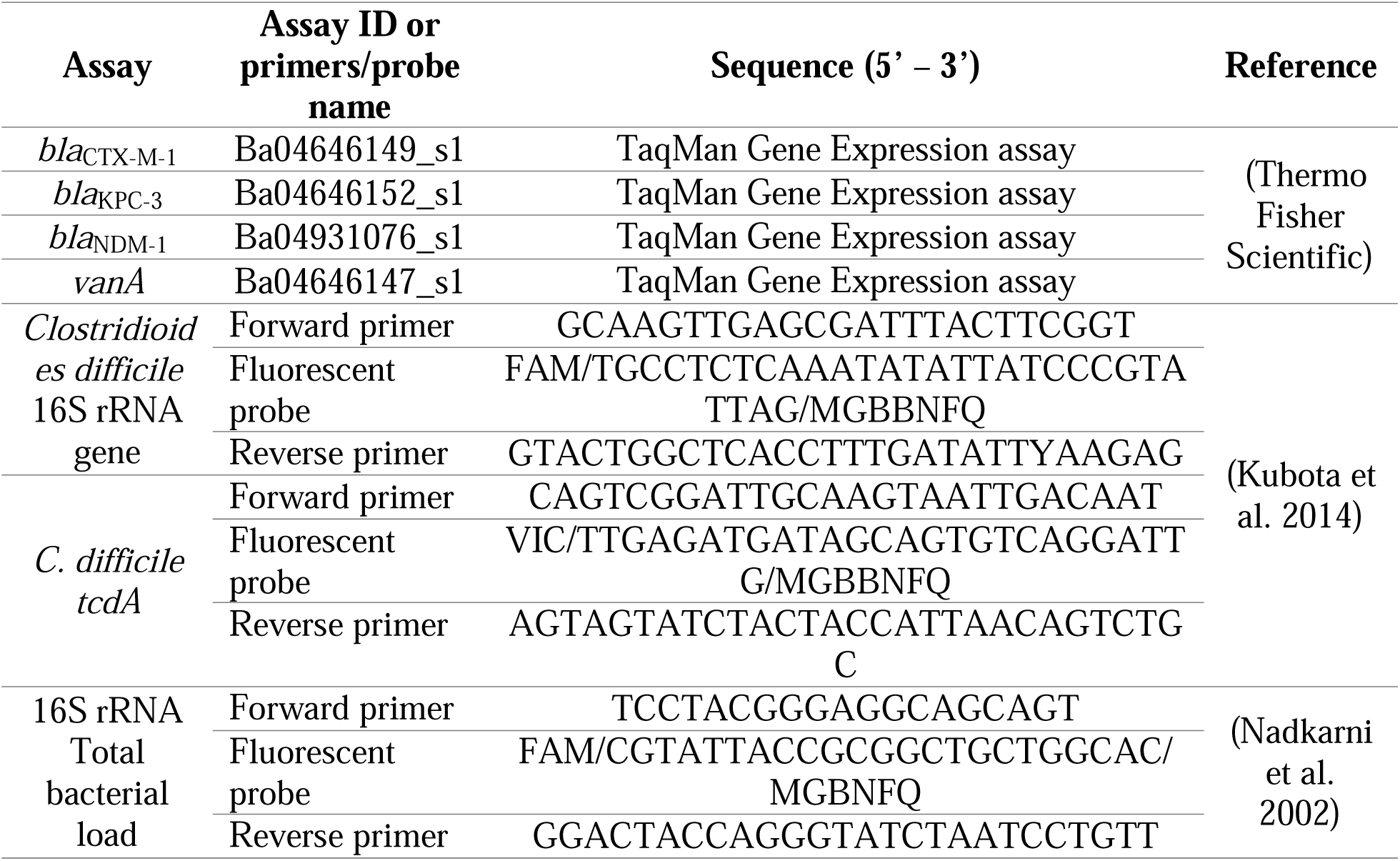
qPCR assays used for ARGs and total bacterial load quantification of wastewater samples.

**Table 3S.**
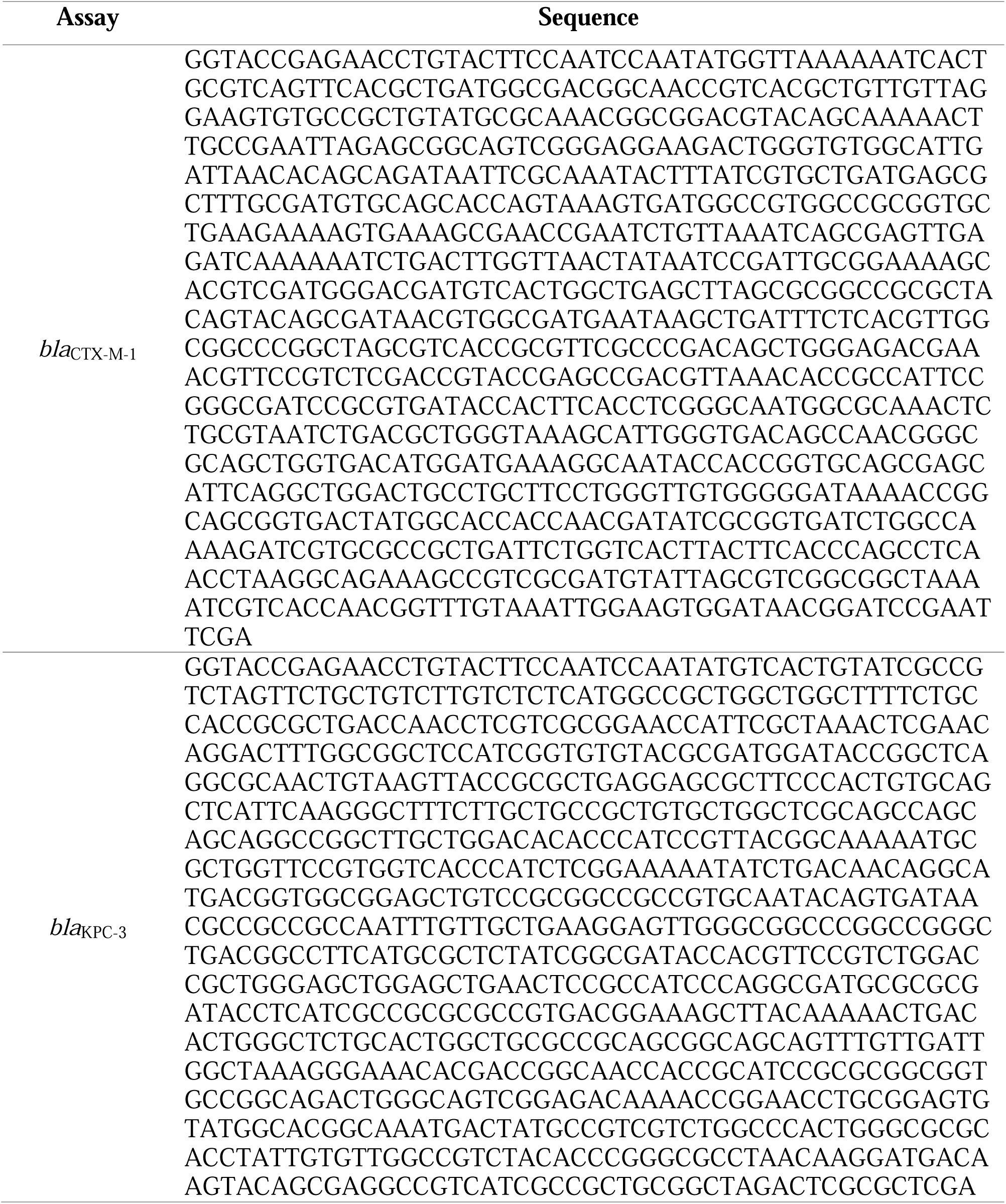
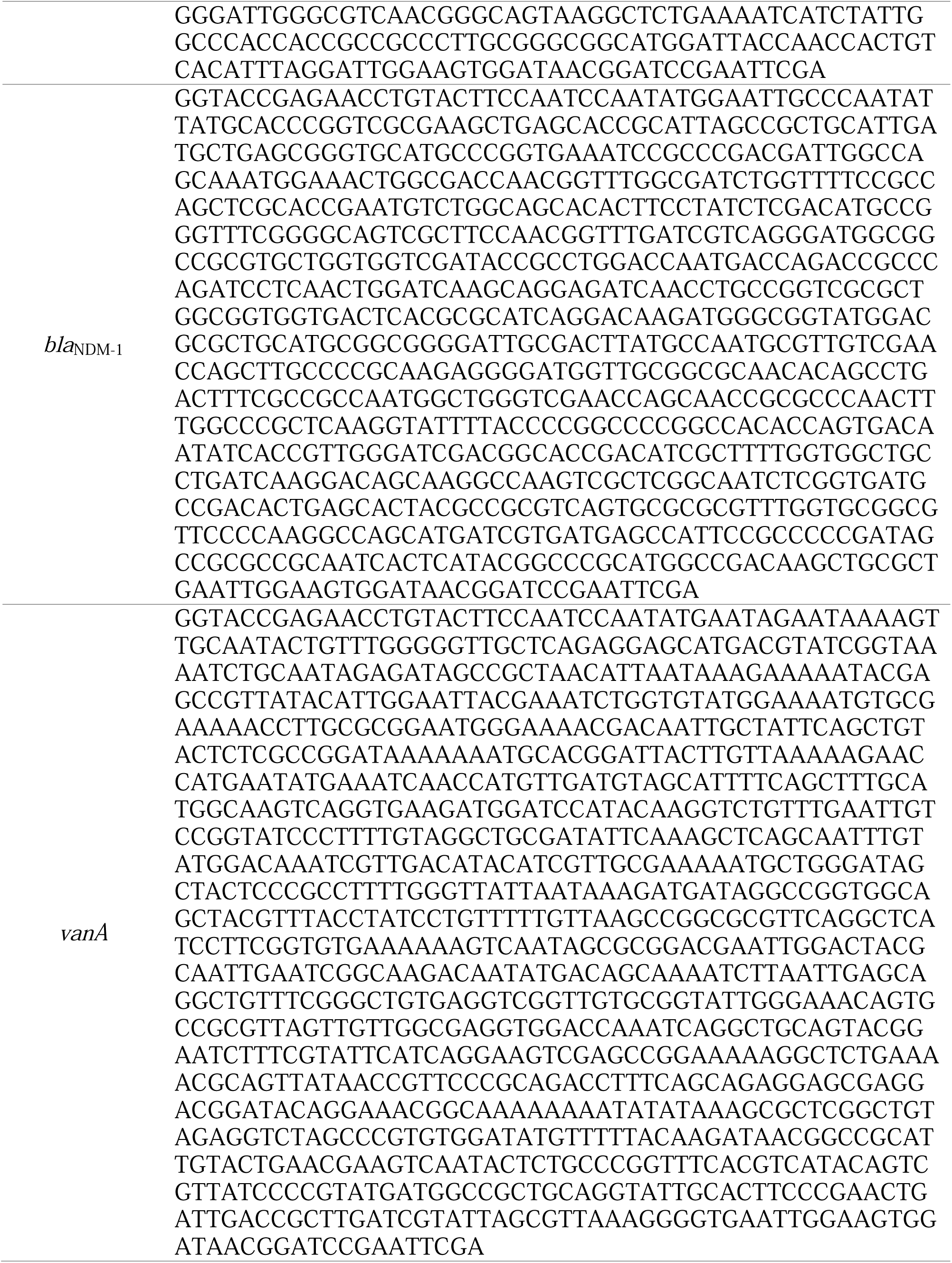
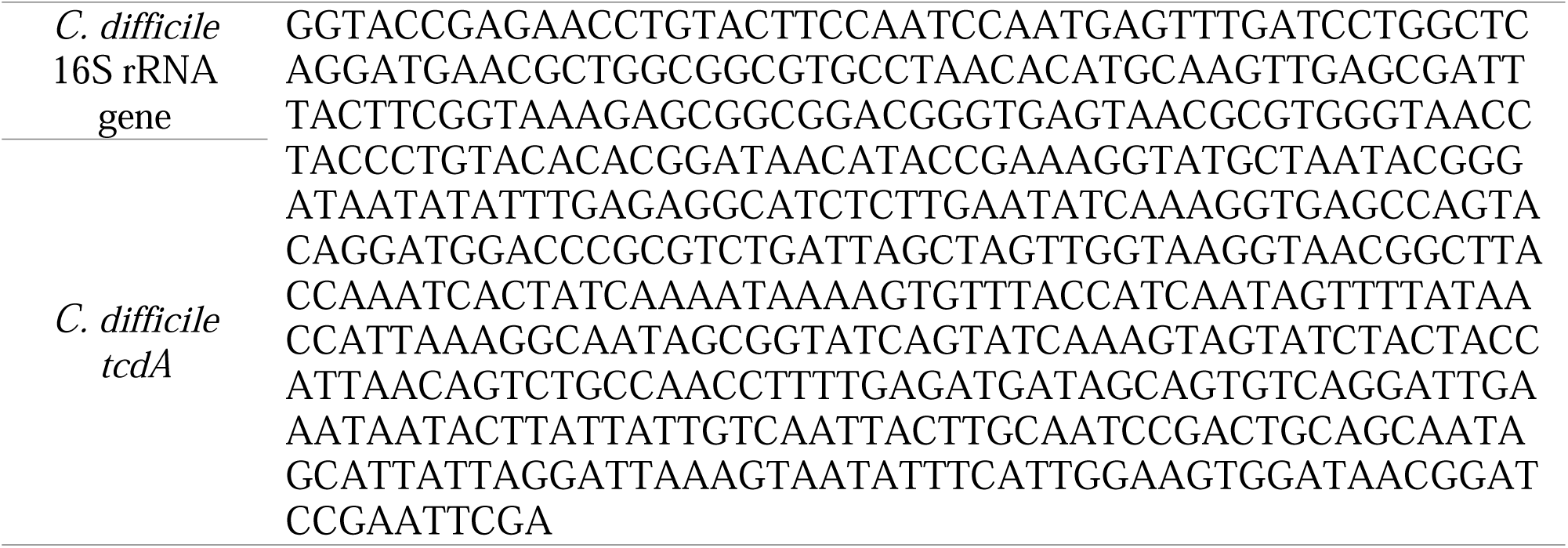
Double-stranded DNA fragments (gBlocks) used as standard curve for qPCR analysis.

**Table 4S.** Number of 16S rRNA normalized aligned reads to each antimicrobial resistance gene at ARG-subtype level. Attached excel file.

## SUPPLEMENTARY FIGURES

**Figure 1S.**
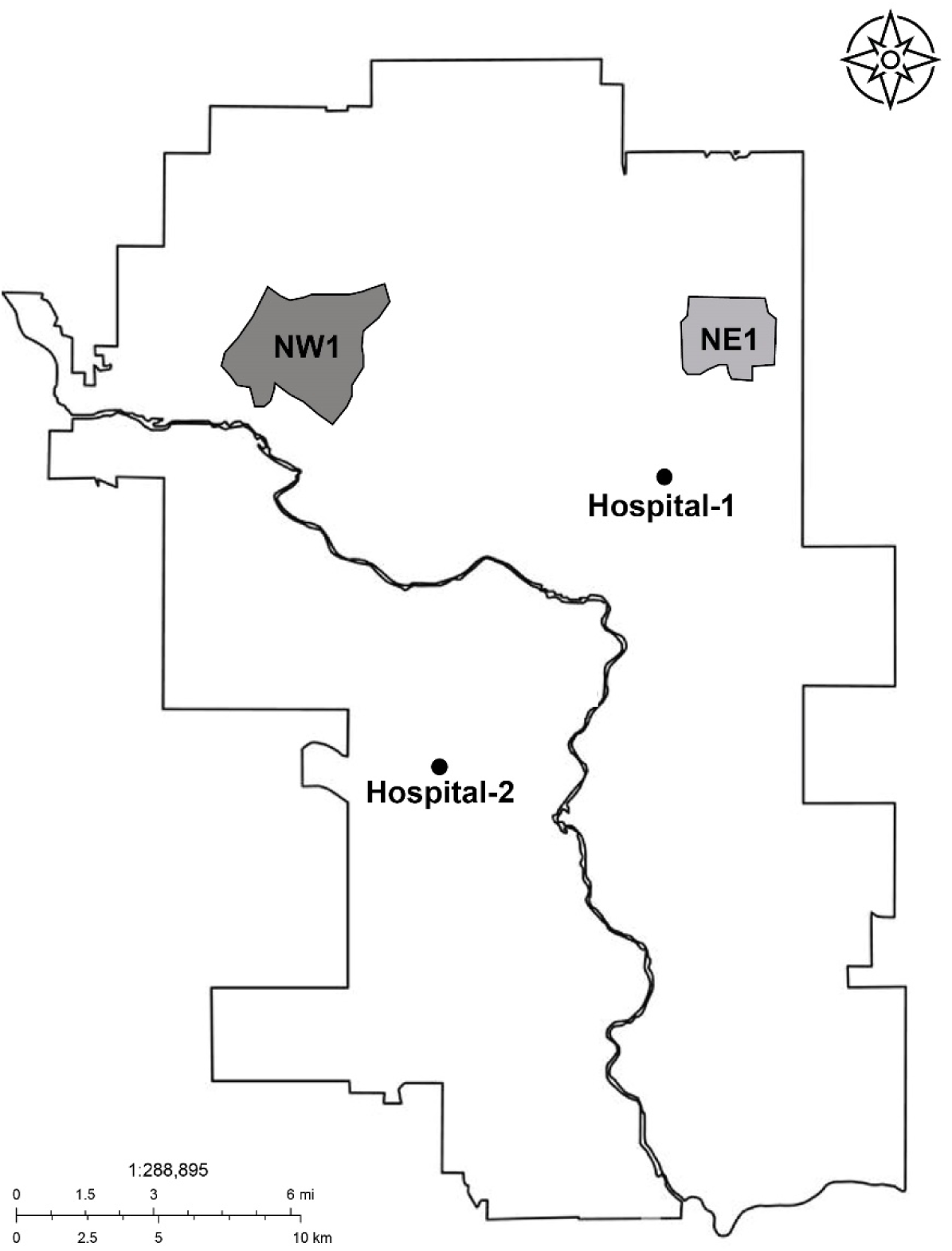
Map of Calgary showing the two hospitals and the two targeted neighborhoods included in the study. Hospital locations are indicted as dots and neighborhoods’ catchment area are shown by the smaller shaded regions and are designated by quadrant: NE1 and NW1.

**Figure 2S.**
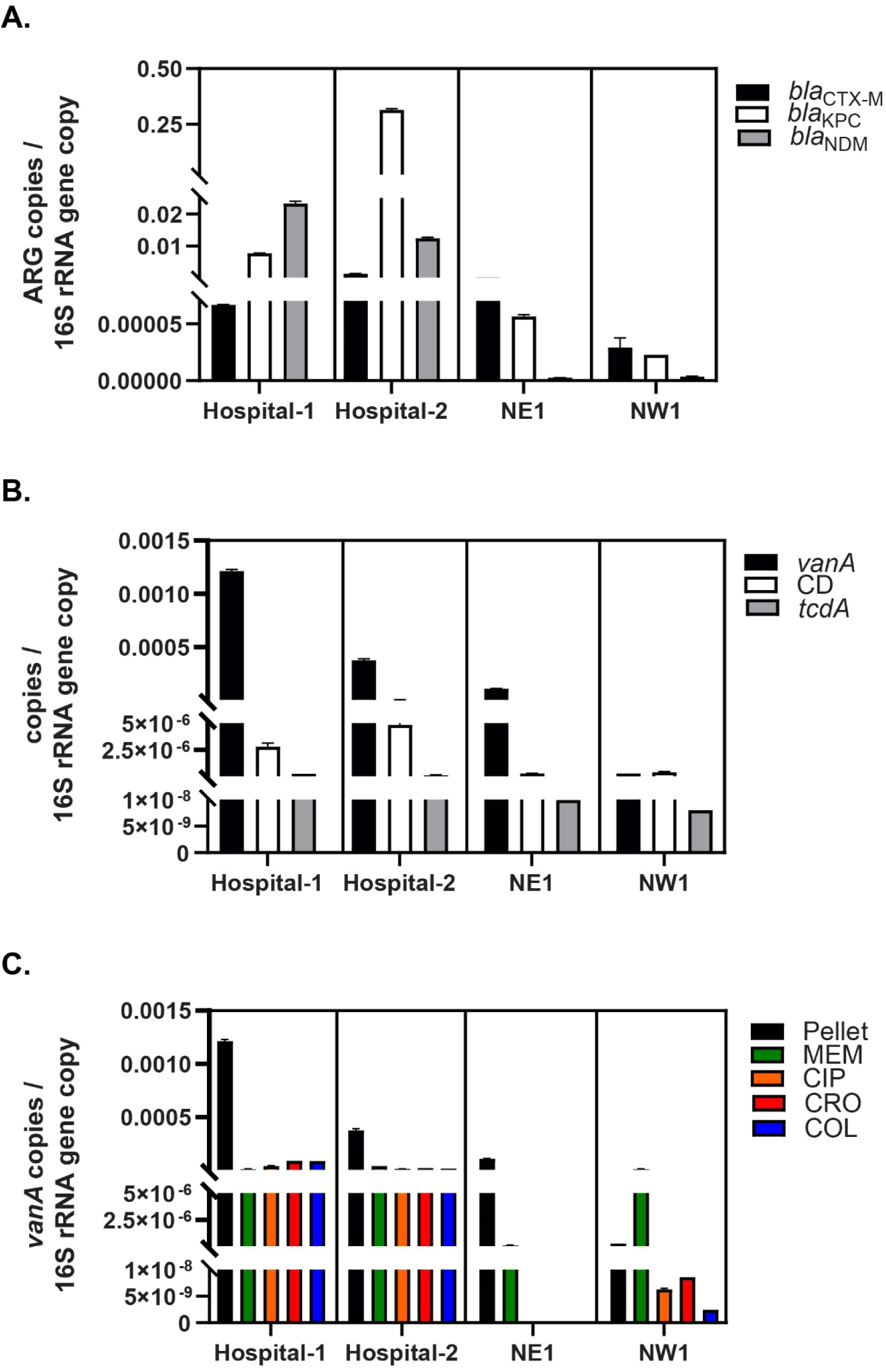
Antibiotic resistance gene determinants. **A.** Absolute quantification of targeted ß-lactamases normalized by the total bacterial load in raw wastewater pellet samples collected from hospitals (Hospital-1 and Hospital-2) and neighborhoods (NE1 and NW1) locations. **B.** Total burden of targeted antimicrobial resistant organisms (AROs)-related gene determinants (i.e., Vancomycin-resistant Enterococcus (VRE): *vanA* and *Clostridioides difficile* (CD)) normalized by the total bacterial load in raw wastewater pellet samples. **C.** Absolute quantification of *vanA* normalized by the total bacterial load in raw wastewater pellet and culture-enriched pellet samples (i.e., MEM: meropenem, CIP: ciprofloxacin, COL: colistin and CRO: ceftriaxone).

